# Guidance on the human-centred design of effective work procedures to support healthcare performance: a pragmatic consensus study

**DOI:** 10.1101/2025.11.26.25341049

**Authors:** Paul Bowie, Fran Ives, Sue Hignett, Laura Pickup, Chris Ramsden, Alastair Ross, Tracey Herlihey, Meredith Makeham, Helen Vosper, Mark Sujan, Carl Horsley, Carl de Wet, Andrew Carson-Stevens

**Author notes:** **Corresponding author:** Prof Paul Bowie, Programme Director (Safety & Improvement), NHS Education for Scotland, 102 West Port, Edinburgh, EH3 9DN, United Kingdom X/Twitter: @paulbowieHF LinkedIn: www.linkedin.com/in/paul-bowie-ab3b49a. https://humanfactorseverywhere.com/.

## Abstract

**Background:** The inadequate design of work procedures is often cited as a contributory factor in patient safety incidents globally, indicating a potential learning need for healthcare teams. Further, the recent COVID-19 pandemic has necessarily witnessed a proliferation of new and re-designed work procedures introduced as staff adapted to altered working conditions, work environments and expanded job roles. In response, this study aimed to rapidly develop guidance on the human-centred design of work procedures by and for healthcare teams.

**Methods:** A rapid, pragmatic consensus building study using a modified-Delphi involving international multi-professional ‘experts’ was undertaken during April 2020. 66 study participants comprising healthcare professionals (n=49) and Human Factors/Safety Science specialists (n=17), based across six countries, were identified from healthcare networks and contributed to the rapid guidance development, informed by Human Factors design principles.

**Results:** Ten key guidance steps and descriptors were identified and agreed upon on how to better develop and implement work procedures based on Human Factors design principles. Examples include: ‘Ensure a procedure is needed’; ‘Involve the whole team’; ‘Identify the hazards’; ‘Capture work-as-done’; ‘Test it out’; ‘Train people’; ‘Put it into practice’ and ‘Keep it under review’.

**Conclusions:** The developed guidance outlines a series of ‘good practice’ principles for care teams in this area. The guidance is freely accessible and will be of interest to healthcare teams at the ‘sharp end’ of clinical practice, risk, safety and improvement advisors, and educators at all levels internationally both in response to crises situations and in routine healthcare delivery.

## INTRODUCTION

At the outset of the Covid-19 pandemic [1], healthcare organisations and teams worldwide were required to respond and adapt rapidly to new ways of working to cope with and manage this unprecedented, dynamic and complex situation. Care teams were extending or adjusting their roles to deliver care in unfamiliar settings with unfamiliar equipment. New or redesigned care procedures were also created to support safe performance now and in the future as health systems transitioned to a ‘new normal’ in how everyday care work is delivered [2].

Work procedures such as written instructions, checklists, and decision-aids are essential tools in supporting safe and efficient team performance in healthcare systems worldwide [3]. One definition of a procedure is “… *an agreed safe way of doing things…usually consisting of step-by-step instructions and related information needed to help carry out tasks safely…”* [4]. Their use is routine in almost every facet of care service delivery to support the performance of important, often safety-critical, work tasks [5]. Typical examples of where they are required to support care teams include: individual and team handovers; donning, doffing and disposal of personal protective equipment (PPE); ordering laboratory tests; triaging patients; and using medical devices.

However, the effective design and implementation of work procedures can be problematic for care teams [6]. Procedure ‘non-adherence’ is often cited as a contributory factor following patient safety incidents [7–8]. Too often, we see examples of where procedures are not needed, are infrequently or never used, are difficult to follow or find, or are unrepresentative of how care teams really perform the work involved [9–11]. Evidence highlights that multiple usability issues are often associated with these performance support tools [12–13]. Furthermore, the topic does not appear to feature on most healthcare training curricula, despite it being a key practical skill and intervention in support of patient safety and other organisational outcomes (e.g. productivity, efficiency, human wellbeing). This highlights a potential learning need for healthcare teams on how to successfully develop and implement work procedures that are usable, useful, and sustained in practice.

Their creation by care teams would therefore benefit from informed ‘good practice’ guidance on user-centred design, implementation and evaluation all of which are vital to sustainable use and successful impact in local practice. For the United Kingdom (UK) Chartered Institute of Ergonomics and Human Factors and NHS Education for Scotland (CIEHF/NES, Box 1) the stimulus for this guidance was the early opportunity to rapidly and freely share expertise and resources by working in close partnership with clinicians and others to support healthcare performance during this crisis period. As a science discipline and applied practice, Human Factors/Ergonomics (HFE) is primarily concerned with the evaluation, (re)design and improvement of human work (Box 2) and is, therefore, well-placed to advise and support care organisations and teams with work transitions and re-designs [14–16]. Given the known problems with work procedure design and the lack of tailored guidance for care teams, it was agreed that this was a logical first step as this was clearly an area where the HFE contribution could help address this specific issue at this time.

This paper describes the process and outputs of this rapidly convened project led by the CIEHF in partnership with NES and other healthcare organisations. The aim was to bring together ‘experts’ in frontline care work and Human Factors specialists to jointly develop and publish advisory guidance on the user-centred design of work procedures by and for healthcare teams. It was anticipated that the target audience for promotion and use of the guidance in response to COVID-19, other crises situations and in everyday ‘normal’ practice would include:

1. Any frontline care team to immediately support rapid work redesigns, task transformation and standardisation as necessary responses to the ramifications of the pandemic on service delivery.
2. Organisational risk, safety, infection control and quality improvement advisors to raise awareness of the issue and this resource, and for them to adapt locally as a teaching tool for care teams.
3. Clinical educators to highlight this as a workforce learning need and provide sufficient information and teaching material to integrate within existing training curricula.

## METHODS

### Study timescale, setting and participants

Over a 3-week period in April 2020, the CIEHF special interest group (SIG) for healthcare assembled, led and co-ordinated a volunteer, online development group (ODG) of informed clinicians, Human Factors specialists and patient safety advisors based in the United Kingdom (UK) and internationally to engage in the rapid development of the guidance on work procedure design. A total of 66 study participants (49 frontline healthcare staff and 17 human factors specialists), representing 13 professional groups based across six countries, were identified from healthcare networks and contributed to the rapid guidance development process (Table 1).

**Table 1.** Characteristics of Online Development Group Members (n=66)

| **Characteristics** | **n** |
| --- | --- |
| ***Professional Groups (n=13):*** |  |
| Human Factors/Ergonomics Specialist (CIEHF Registered Members) | 17 |
| Primary Care Clinician | 17 |
| Secondary Care Clinician | 18 |
| Clinical Engineering Lead | 2 |
| Executive Leader | 2 |
| Quality Improvement Advisor | 2 |
| Ambulance Paramedic | 2 |
| Clinical Risk Advisor | 1 |
| Healthcare Administrator | 1 |
| National Clinical Skills / Simulation Leader | 1 |
| National Patient Safety Leader | 1 |
| Patient Safety Lead Educator | 1 |
| Primary Care Manager | 1 |
| ***Base Country of Participants (n=6)*** |  |
| England | 27 |
| Scotland | 22 |
| Australia | 11 |
| Wales | 4 |
| USA | 1 |
| New Zealand | 1 |
| ***Gender*** |  |
| Male | 39 |
| Female | 27 |

### Human Factors Design Principles

Guidance development was informed by well-established good practice principles for user-centred design - a sub-branch of Human Factors science and a process for ensuring that the needs, wants, preference, capabilities and limitations of people (i.e. guidance users) are the priority focus throughout every development stage [17–20]. User application of these principles in the local design of work procedures potentially helps to ensure that they are necessary, usable, valued, reflective of work reality, and contribute to reducing task-related risks.

### Guidance development process

The virtual development process was aided initially by a rapid literature search and identification of key source material to draw upon. This evidence was augmented by accessing and applying the ‘expert’ knowledge and experience of user-centred design principles to the development process by suitably qualified Human Factors specialists in the ODG. Most of whom had significant workplace experience of this issue in high-risk industries, including regularly training healthcare teams on procedure, protocol and guideline design and implementation. The ODG used Microsoft Teams and electronic mail to quickly co-ordinate, communicate, and capture feedback from group participants, and then build consensus on the written content and interactive graphic design of the guidance documents over the short timescale involved.

### Pragmatic Consensus Agreement

The approach to building consensus was informed by the modified-Delphi method [21]. After three Microsoft Teams meetings and multiple email exchanges (with all participants attending at least a single meeting or responding to emails) the initial guidance document was drafted by the two study leads (PB and FI), experienced in consensus building methods and patient safety research. This was subsequently emailed to all ODG members for critical review, comment and feedback. This process was repeated three times. The study leads filtered and incorporated all feedback (where judged useful) for each iteration. All group members commented on at least one occasion during this content development and consensus building process. It was pragmatically agreed that consensus was reached on the fourth iteration when no new significant feedback was received from the group, other than correction of minor grammatical and typographical issues.

## RESULTS

### Summary of Key Guidance Steps

The ten key guidance steps identified and agreed on how to design work procedures better are summarised as follows (Figure 1):

**Figure 1.**
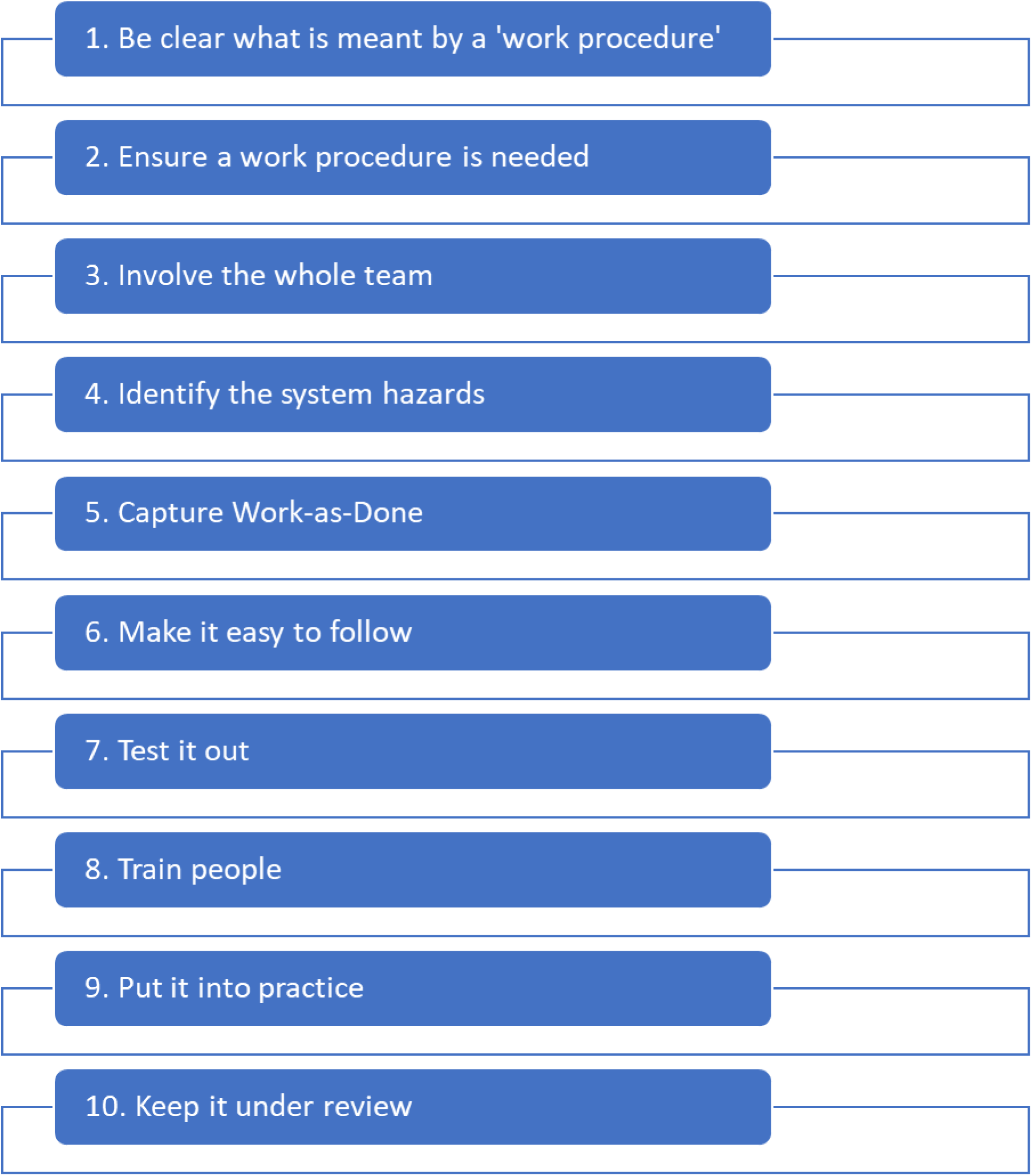
Ten Steps: Human-Centred of Healthcare Work Procedures.

#### Step 1 – Be clear about what is meant by a ‘work procedure’

It is important to understand what is meant by a ‘work procedure’ and to differentiate it from other related tools (Box 3). A procedure is simply defined as a logical step-by-step way of doing things at work, often in the form of written instructions, checklists, decision aids, diagrams, or flow charts [3–4]. Sometimes they are known as standard operating procedures (SOPs) or maintenance procedures. Work procedures are useful tools for care teams because they are a means of, for example: agreeing and standardising how things are done; clarifying responsibility (i.e. who is responsible for which steps); ensuring recommended good practice is followed; and reducing risks to as low-as-reasonably-practical (ALARP – [22]) for patients, team members, our organisations and others.

#### Step 2 - Ensure a work procedure is needed

Care teams should think carefully about whether a procedure will be needed and if it will be used. Good examples of where procedures are important include safety-critical, complex or important tasks that need very clear instruction, or when compliance with standards, good practice or legislation is required [23]. Once there is agreement amongst the team, it should be clearly understood that the procedure alone will not solve the problem and that team ‘buy-in’ and co-design are vital to successful and sustainable implementation.

#### Step 3 - Involve the whole team

Those leading the design of a procedure should be closely involved in the related work and will be a procedure user, as it is important to have a practical, realistic understanding of the tasks at hand. The views of leaders, managers and supervisors are important but appreciate that their perspectives on how the work is really done could be out-of-date or unrealistic. A full understanding is needed of who will be using the procedure and when it is needed. All relevant team members, including patients, clients, carers and families (where appropriate), should be involved in the design, implementation and review of the procedure at every stage [24–25]. In practical terms, this means understanding the wider system of which the procedure is a part. This applies throughout this guidance, but particularly for Steps 3, 4 and 5. There are several methods available to support this including, Systems Thinking for Everyday Work (STEW) discussion cards (26); systems mapping workshops [27]; and the Systems Engineering Initiative for Patient Safety (SEIPS) [17] which is described in Step 4.

#### Step 4 - Identify the hazards

Explore what the worst things that can happen are and how they might occur [28–30]. To help develop the procedure more effectively, all members of the team need to think through and identify all the things that can possibly go wrong (hazards) using a human factors systems approach (like those described in Step 3) to guide this process by considering:

`o` *People issues* e.g. the complexity of the patient’s or service user’s condition and their needs; the physical, emotional and mental limitations and capabilities of staff
`o` *Job tasks* e.g. how complex, hazardous and demanding are these?
`o` *Tools and technology* e.g. the availability, accessibility, usability, safety, mobility and positioning of equipment.
`o` *Physical environment* e.g. the location, size, layout, noise, lighting, temperature and design issues related to the working environment.
`o` *How work is led and organised* e.g. influence of rotas, staffing levels, leadership styles, and local culture.
`o` *External influences* such as national policies, targets or regulations.

#### Step 5 - Capture work-as-done

If procedures are to be useful then they need to accurately reflect ‘work-as-done’ - the everyday realities experienced by those on the local frontline, rather than ‘work-as-imagined’ by those at a distance from the frontline who do not currently do or understand the job [31]. ‘Work-as-done’ will look different depending on the diverse and variable work conditions the care team faces such as when they are very busy, short-staffed, missing equipment or dealing with difficult, uncertain or complex care scenarios. The procedure needs to be designed to take these contexts into account. Observe and speak to people as they perform tasks related to the procedure. Are there different ways to carry out the procedure depending on variable work conditions? How do people cope with the competing work priorities they face?

#### Step 6 - Make it easy to follow

How the procedure is visually designed and set out is essential to people engaging with and using it without difficulty or frustration. The goal is to make the procedure concise, workable, relevant, realistic and clear. Overly long procedures should be avoided to optimise user attention – ‘less is more’. To do this:

- use plain language and agreed terms that everyone can understand;
- keep it short and as simple, less than 15 words per sentence;
- avoid large chunks of texts. If they are necessary, break them up with diagrams or flowcharts;
- write using an active voice (“place label on the specimen container”) and not in a passive voice (“the specimen container should be labelled”);
- and increase readability and emphasise key parts of the procedure (e.g. use consistent font types and sizes that are easy to read and follow such as Arial size 11 or larger)

Remember that if everything is emphasised then nothing is emphasised. There are number of resources available to support users in understanding practical information presentation [32–36].

#### Step 7 - Test it out

Ask the people who will use it to review the draft and provide feedback on improving it. Get feedback from people with a range of experiences including those who have never done the work before, and those who were not involved in the design of the procedure. Do this repeatedly until everyone is satisfied with it. Test the procedure in ‘real-life’, while observing and discussing its use. Use plan-do-study-act (PDSA) cycles to help with review and improvement [37].

#### Step 8 – Train people

People need to be trained in the use of the procedure. Identify how formal training needs to be and the most effective way such as face-to-face training, online packages or discussion during a team briefing. Identify who can best deliver the training. An appropriate person-in-charge, e.g. manager for the area, should ensure that all relevant staff have received training, including scheduling refresher training. Collect feedback from the training sessions to use for future improvements of the procedure.

#### Step 9 – Put it into practice

Care teams need to ensure the procedure is used. This requires as much effort, if not more, than producing it. Simply emailing staff to inform them, or putting up a poster, will have limited or no impact on them remembering to use the procedure at the right time. If you have involved the team throughout the design process, then these issues will be less challenging. Consider the following:

- Where it is stored so that it is easy to find (electronic and hard copy)
- How to communicate the location of the procedure; the best format (electronic and hard copy); and the name of the procedure. Is it logical?
- Make sure there are measures in place to educate new staff e.g. as part of induction processes.
- All versions should be dated and have an identifiable name and issue number, previous versions should be removed to eliminate confusion and the risk of using out-of-date documentation.
- Hard copies of the procedure should be in good condition, not dirty, torn or with pages missing. If they are for use in a clinical area, they will need to be laminated to facilitate cleaning.

#### Step 10 - Keep it under review

If a procedure is not being followed, positively explore and understand any ‘deviations’. Learning through incident reporting and team-based event reviews can explore how or why procedures may not be fit for purpose. Schedule regular reviews of procedures, e.g. weekly, monthly, yearly. This will ensure the procedure matches changing ‘work-as-done’. Monitor gaps between procedures and the way ‘work-as-done’ occurs. This will alter with new equipment, changes in staffing levels, an incident, new guidelines and changes in the environment. A growing gap often indicates that the procedure needs updating. Withdraw or redraft a procedure if the desired effect is not being realised.

## DISCUSSION

The rapid development project met its aims in agreeing and setting out a 10-step guiding process for care teams to methodically develop better work procedures based on existing good practice. To deal with the challenges of the Covid-19 pandemic, healthcare teams needed to adapt in agile ways to peaks in demand, working patterns being disrupted, workforce absences due to illness, and restrictions in the way care is delivered to safeguard patients and staff [2]. Suitably well-designed work procedures are an integral part of doing this safely and efficiently [4,6,29].

The guidance is available freely to care teams and educators in every sector worldwide to help improve the design and refinement of their work procedures. Early examples of where it has been quickly applied include in the design of bedside guides for routine tracheostomy care and ventilator emergency care for Covid-19 patients [38], and in risk assessment processes in Scottish primary care facilities [39]. The guidance can potentially support current and future working practices and contribute to, for example, ongoing patient safety, quality improvement, organisational learning and workforce wellbeing initiatives. It is also of high relevance to care educators, and those in training positions. Gaining experience of leading the effective co-design, testing, implementation and review of work procedures needs to be recognised as an important professional skill in preparation for everyday health and social care practice.

The developed guidance is arguably robust enough given the extent and scale of contributions from ‘experts’ in the time available who are informed on the topic. Most contributors are experienced leaders in their respective fields and have previously published in relevant areas e.g. patient safety, healthcare human factors, risk, and improvement. While the development process was informed by multidisciplinary contributions and a consensus building process, the context of Covid-19 has meant a trade-off between thoroughness and efficiency was inevitable because of the rapid process that was employed (40]. It would have benefited from contributions of more diverse and representative groups of target users, and a more rigorous review of existing evidence. In an ideal world, procedure development by care teams should be informed by rigorous and systematic studies of work-as-done, supported by appropriate Human Factors methods e.g. hierarchical task analysis [41] and a prospective hazard assessment process [42]. However, knowledge of these is limited in the care workforces. Finally, making available accessible guidance on the user-centred design of work procedures is only the first step in trying to achieve the goal of making work procedures more usable and effective. Like any complex intervention, the implementation uptake and impact of the guidance, in both educational and frontline care settings, will require significant study to better inform the contexts and mechanisms underpinning if and how it ‘works’, where, when and for whom [43].

## CONCLUSION

The developed guidance outlines a series of principles for care teams that are conducive to good practice in this area. In summary, for a work procedure to be fully accepted and used, the relevant care team should be involved from the start and throughout the design process. Team members must all agree that it is needed to support work performance and then actively participate in its development and review – ultimately, they must believe that its use would be better than what is currently being done. Without team involvement or agreement, it will be difficult to properly develop, introduce and sustain a new procedure when it is most needed in routine or rare work situations [44].

To access the full guidance on the design of work procedures with illustrated care examples, please visit: https://covid19.ergonomics.org.uk/

## Data Availability

All data produced in the present work are contained in the manuscript

## Acknowledgments

Noorzaman Rashid (Chartered Institute of Ergonomics and Human Factors, UK); Peter McCulloch (University of Oxford); Duncan McNab, Sarah Luty, Anna Alexander, Lynne Innes, Tracey Cricket, Mark Johnston, Jean Ker (NHS Education for Scotland, UK); Giulia Miles, Evi Burford, Bryn Baxendale (Nottingham University Hospitals, UK); Michael Moneypenny (Scottish Centre for Clinical Simulation and Human Factors/NHS Forth Valley, UK); Alastair Williamson (University Hospitals Birmingham NHS Foundation Trust, UK); Dave Murray (James Cook University Hospital/Health Education England, UK); Haroon Ahmed, Adrian Edwards (University of Cardiff, UK); Donna Forsyth (NHS England and NHS Improvement, UK); Graham Forman, Gary Rutherford (Scottish Ambulance Service, UK); Carl Horsley (Middlemore Hospital, New Zealand); Thomas Jun (Loughborough University, UK); Courtney Grant (Transport for London, UK); Samantha Fell (Spectrum Lakeland Health, USA); Peter Ledwith (Advancing Quality Alliance, UK); Nathan Anderson (Doctor-in-Training, University of Cardiff, UK); Melanie Newton (Teesside University, UK); Kim Hinshaw, (Sunderland Royal Hospital, UK); Heather Gallie (Salford Royal Foundation Trust, UK); Julie Hannah, Wendy Russell (NHS Ayrshire and Arran); Malcolm Philips, Scott Clarke, Ed James, Simon Paterson-Brown (NHS Lothian); Ted Mullen (NHS Greater Glasgow and Clyde, UK); Manoj Kumar (NHS Grampian); Nikki Davey, John Pickles, Martin Bromiley (Clinical Human Factors Group); Thomas Rollinson (East Midlands Ambulance Service NHS Trust, UK); Huw Gibson (Rail Safety and Standards Board, UK); Sarah Tapley (Chartered Ergonomist and Human Factors specialist, UK); Jane Higgs (CIEHF, UK); Luke Worth, Rolando Villegas, James Lind, Rebecca Fawcett, Nikia Goldsmith, Ashlea Walker, Kahlod Ali, Ian Scott, Kathryn Thompson, Simon Bugden (Queensland Health, Australia), Janette Edmonds, (The Keil Centre Limited, UK).

## Declaration of conflicting interests

The Author(s) declare(s) that there is no conflict of interest.

## Contributorship Statement

Study conception and design: P. Bowie and F. Ives. Data analysis and writing: P. Bowie, F. Ives, H. Vosper and A Carson-Stevens. All authors were participants in the online development group and provided feedback on the written manuscript as well as making suggestions for additional resources signposted within this document.

## Funding with award/grant number

NHS Education for Scotland, Edinburgh, UK

## Ethics

Not required. This type of study is exempt from ethical review in the UK as it is classified as service evaluation rather than research.

## Box 1.

**About the CIEHF and NES**

- The CIEHF is the United Kingdom (UK) based professional society and regulator for ergonomists and human factors specialists, and those involved in user-centred design
- In response to the Covid-19 pandemic, the CIEHF set up expert panels to advise and support related health and social care activities and wider society: https://covid19.ergonomics.org.uk
- NES is an education and training body and a national health board within NHS Scotland. It is responsible for developing and delivering healthcare education and training for the NHS, health and social care sector and other public bodies. It has Scotland-wide role in undergraduate, postgraduate and continuing professional development. The NES Safety and Improvement Research Team led and co-ordinated this development work in partnership with CIEHF.

## Box 2.

**About Human Factors/Ergonomics**

- “Ergonomics (or human factors) is the scientific discipline concerned with understanding the interactions among humans and other elements of a system, and the profession that applies theory, principles, data, and methods to design in order to optimise human well-being and overall system performance” (International Ergonomics Association: https://iea.cc/)
- HFE emerged as a scientific discipline in the 1940s out of the growing realisation that, as technical equipment and work system designs became increasingly complex, not all of the expected benefits would be delivered if people were unable to understand and use the equipment or interact with a system to its full potential.
- Application of HFE principles and methods is well established in many complex and safety-critical industries such as those found in the energy, transport and defence sectors
- While not well-established HFE approaches in healthcare have informed incident investigation processes, safety culture measurement, ambulance vehicle design, clinical education, medical device usability, complex system analysis, risk assessment and control, work system safety, PPE design, quality improvement approaches and the built environment.
- Overall, despite increasing recognition of its importance in improving organisational performance, patient safety and workforce wellbeing, the discipline has had limited impact in health and social care due to lack of policy attention and resource investment

## Box 3.

**Basic definitions of work procedures and related tools**

- **Guidelines** (evidence-based good practice statements)
- **Policies** (outline guiding principles),
- **Protocols** (defines procedures to be followed)
- **Work Procedures** (step-by-step instructions on how to carry out a task)

## Notes

### Competing Interest Statement

No competing interests declared

### Funding Statement

This study did not receive any funding

